# Liver Echinococcosis Lesion Classification Tool by Deep Learning: development, deployment, and validations

**DOI:** 10.1101/2022.01.27.22269985

**Authors:** Liang Huang, Xiaohong Xi, Zhu Chen, Yilan Zeng

## Abstract

The endemic of Echinococcosis imposed heavy disease burden in some areas. The sonography for Echinococcosis lesions was essential to disease diagnosis and managements. Especially the biological typing of lesions was key to disease treatments. We used deep-learning tools to help sonographer to classify the lesion types. The model achieved 85%(302/376) accuracy, in contrast to senior sonographer achieved 72%(61/85) accuracy. The accuracy of AI model was higher than senior sonographer (p-value=0.01), could be a feasible method to help sonographer in remote area.

## Introduction

Echinococcosis is a result of human infection by larval stages of taeniid cestodes of the genus Echinococcus. Echinococcus *granulosus* causes cystic echinococcosis(CE), Echinococcus *multilocularis* causes alveolar echinococcosis (AE), two separately diseases with different biological processes^1,2^. The co-infection of the two pathogens was rare^3^. At least 58% of the population are at risk of acquiring echinococcosis, distributed in Europe, North and Central America, Africa, Australia and Western China^4^. It is a potential fatal zoonosis^5^, a major public health concern in Qinghai-Tibet Plateau due to the highly pandemic of both CE and AE. The echinococcosis DALYs in the Tibetan communities were 126,159 life years annually estimated by Wang^6^. The Tibet population in China holds 1.66% prevalence of both CE and AE, estimated 50 million people are at risk of infecting the disease in Western China^7^. China has been investing large amount of resources to control echinococcosis, launched national control program^8^. The strategies of the national control program majorly included definitive hosts deworming, slaughtering control, and intermediate hosts vacation^2^. Ultrasound screening was included as an essential measure to find the patients and follow-up.

Echinococcosis is chronic parasites disease, liver echinococcosis were most common in clinical^9^. The treatments for it included surgically remove the lesion, PAIR(puncture, aspiration, injection, and respiration), albendazole or they combined^10,2^. The lesion classification for CE is the one of the key aspects in patients ‘ management, since the classification was closely corelated to the underlying biological status of the larva.

For CE, the ultrasonic WHO classification is commonly used (CL, CE1, CE2, CE3, CE4, and CE5). The CE1-2 types of CE indicating active infection should be treated by Albendazole or/and surgical resection. The CE4-5 are considered degenerated lesion due to failure of natural infection or effects of treatments, should be closely monitered^11,12^. The CE3 is considered the transitional phase of lesion. The CE1-5 are the continuous progress of “active-transitional-inactive”^13^ indicating importance for biological process and disease assessments.

The AE is a silent progressing infiltrative proliferation of the parasite, mimicking a malignancy^14^. For AE cases, the lesion classification are more complex and uncertain, research revealed that the clinical stage of AE was not corelated to ultrasound appearance^15^. WHO had proposed a PNM classification system for AE^16^. The PNM classification system (P=parasitic mass in the liver, N=involvement of neighboring organs, and M=metastasis) was designed to systematic evaluation for surgical purpose majorly. Besides PNM classification system, the Ulm university proposed a classification system for AE^17^, and five types of lesion proposed.

The PNM classification systems required extra examinations other than ultrasound, such as CT or MRI scan. However, the PNM system was not feasible for many occasions. The accessibility of medical resources in Qinghai-Tibet Plateau is incomparable with urban area^18^ due to the high altitude and atrocious weather, the local paramedics are lack of professional training, especially for sonography.

This study purposed to develop deep learning-based application to automatically type lesions for both AE and CE, giving diagnostic assistant for local paramedics. In this study, we used 2820 ultrasound images training our model, and used external data to validate the model. It is the largest echinococcosis ultrasound training set so far and its application for was published on a web for open access.

Previous studies had developed deep learning system for hepatology by using medical images, pathology, clinical and laboratory data, natural languages, etc. They are majorly designed to address liver problems such as liver fibrosis(cirrhosis), liver cancer, portal hypotension, etc^19^. Relatively, less study were aimed to the liver echinococcosis, which is on the list of NZDs^20^ by WHO. Before our study, Xin had built a segmentation and classification networks for echinococcosis, shedding light on potential development in this field. However, it used 160 CT scan images for training, and the classification of echinococcosis were not covered all types of both AE and CE^21^. Comparing with CT images, ultrasound was more widely used in filed echinococcosis diagnosis. In this study, we used 2820 ultrasound images for training covering all echinococcosis types.

## Method and Material

### Patient Data

Sonographies were retrospectively obtained from National Control Program for Echinococcosis Screening in Qinghai-Tibet Plateau^22^. The medical decisions were not based on results of this study. All the patients were previously diagnosed echinococcosis by systematic evaluations including medical images and serology tests, fulfilled the criteria of echinococcosis diagnosis criteria.

In this study, we collected 3423 images of echinococcosis patients who were enrolled and routinely managed in national echinococcosis control program.

Ethical approval for this retrospective study was obtained from the Ethics Committee of the Chengdu Public Health Clinical Center. The patient consents for inclusion were waived as the retrospective nature of this study and anonymous data use.

### Classification criteria

In this study, we used CL-CE5 classification system for CE, which was recommended by WHO recommendation^13^. We used infiltrating, calcification, and necrotic lesion for AE classification, which was recommended by Chinses Guideline for AE^23^.

For CE lesions, the CL lesion, which was appears “cystic lesion “, which appears no different from simple cyst in liver, could not be confirmed by ultrasound image solely. The CE1 refers to a simple round or oval unilocular cyst with anechoic content and a visible double cystic wall. The CE2 cyst is filled with daughter vesicles. The CE3 cysts includes two stages, CE3a is characterized by the “water-lily “ sign, CE3b represented by floating membranes. The CE4 typically reveals coarse variable hyperechogenic or hypoechogenic echotexture without daughter vesicles. The “ball of wool “ sign, corresponding to the detached endocyst as a hypoechoic folded structure embedded in a hyperechoic matrix, is the key ultrasound sign. The CE5 cysts are partially (with an egg-shell calcified wall) or completely calcified with shadowing^13^.

For AE lesions, in this study, we followed the Chinese Guideline for AE, which was also applied in National Controlling Program in endemic areas. It proposed three ultrasound types of lesions. The infiltrative lesion (AE1) appearing hailstorm pattern, heterogeneously echogenic areas, in many cases scatter calcifications can be seen. The necrosis lesion (AE2), appearing a pseudocystic pattern with an irregular hyperechoic rim. The calcification lesion, appearing solid hyperechoic lesion (AE3).

### Data labelling

Expert who had more than 15 years of echinococcosis ultrasound diagnosis and ultrasound based echinococcosis patients managements had labeled the included ultrasound images by using LabelMe^24^, an open-source image label tool. The lesions in the images were labeled by different color, and normal liver background was labeled as black. The labeled images were automatically cropped into fixed size of input images (512*512 pixels) for model learning and inference.

### Model constructions

The U-net architecture achieves high performance on different biomedical images segmentation applications by using relatively less quantity of training images^25,26^. In this study, we used two separated U-net models. Firstly, we constructed “Classifier “ U-net model which was used generalized multiclass dice loss (GMD). The GMD penalizes the wrong classifications more aggressively and more effective for training if the unbalanced category of training samples used. However, it produces more noise (false positive predictions) produced. Secondly, the identical “Shaper “ U-net using conventional binary dice loss function focusing on lesion shape learning black-white (lesion-background, values 0 and 1) outputs, omitting the lesion class information. It produced more accurate shape inference for lesions. The inference result produced by multiplication of two tensors of the models ‘outputs of the models’ outputs (Figue 1).

**Figure 1.**
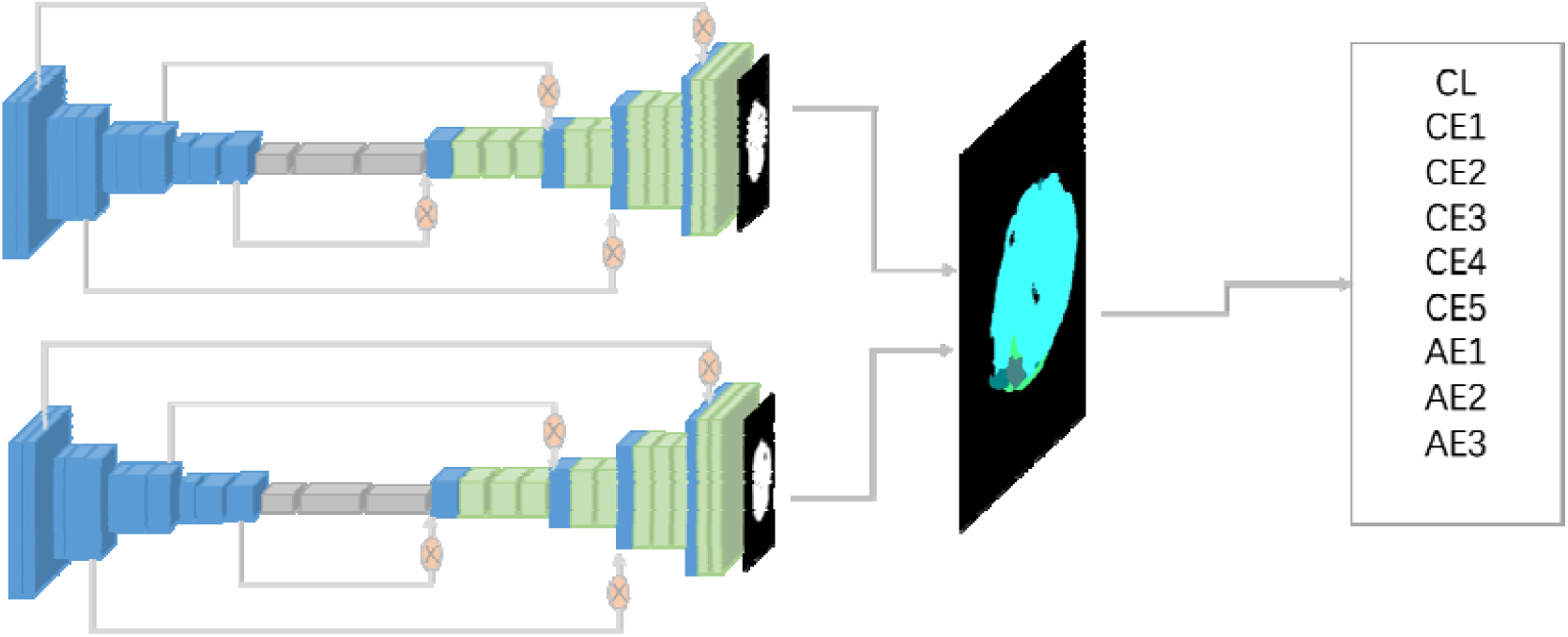
The architecture of Two U-nets model.

The whole program was based on Python programming languages with PyTorch 1.3 deep learning framework. The core construction of the models consist 10 of convectional layers including downsampling, upsampling, and attention gates were used in this model^27^. The models were trained with mini-batch SGD optimizer with momentum on two Nvidia 1080TI GPUs. The learning rate was chosen to minimize the error in the tuning dataset. Early stop training strategies was used for preventing over fitting.

### Web-based user interface

Trained weights files loaded in a web-based user interface under Flask framework for internal logical process and HTML for appearances of the application of both computer and mobile phone. The users will upload images transferred from ultrasound equipment by computer or mobile phone. The web-based UI will automatically calculate the hot-spot of the ultrasound images and crop them into proper size and optimized location(Figure 2).

**Figure 2.**
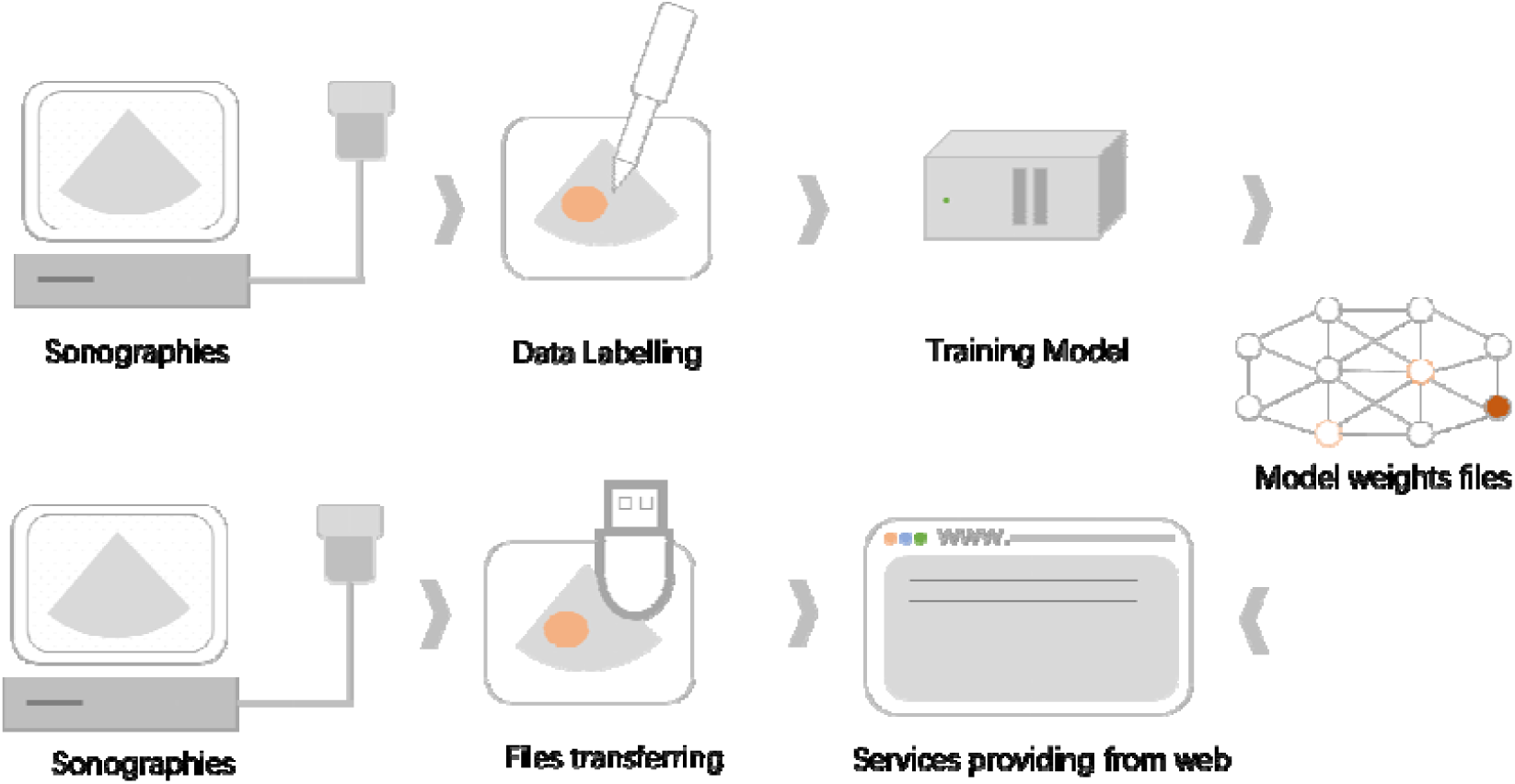
Web Application structure of model

### Model validations

We compared classification accuracies for human and model. The senior sonographer has 15 years of working experiences. The performance of model was designed to achieved equivalent accuracy with senior sonographer.

### Statistical analysis

The statistical analysis was performed with the STATA/SE 14.1 software (StataCorp, 4905 Lakeway Drive, College Station, Texas, USA) and R 4.0.2^28^. The ggplot2^29^ for R was used for diagram plotting. Confusion matrix^30^ was used for validation of the accuracy of the model. The confusion matrix was plotted by ggplot2 packages. The α was set to 0.05. P<α was statistically significant.

### Hardware and software

The sonographies were obtained from various ultrasound equipment, including Mindray™ M5, Seote MyLab™ Alpha. The server used for training model installed two 1080TI GPUs with Ubuntu 18.04 LTS platform. A Nvidia AGX Xavier edge computing device providing web-based interface and model inference. These devices install Ubuntu 18.04 LTS with PyTorch 1.3 and Flask library installed.

## Result

### Dataset

According to our dataset, we had 2250 training images, 190 images validation images and 376 testing images. In the training dataset, for CL and CE1-5 type, 140, 172, 198, 280, and 472 images were trained, respectively. For AE lesion, 420, 290, and 278 images were trained, respectively.

### Training the model and publishing

Model was trained for about 10 hours to achieved convergence after 90 epochs (multiple times of training for optimizing). The generalized dice score of Classifier for validation set was 0.77, the Dice score of Shaper for validation set was, 0.86. The web-based interface uses 3.1s for average for single image analysis in current hardware configuration (each service has six times of inferences).

### Comparison with human sonographer

Total of 376 samples were inputted into AI model, archived 85%(302/376) accuracy. In contrast, total of 85 samples were classified by senior sonographer achieved 72%(61/85) accuracy. The accuracy of AI model was higher than senior sonographer (p-value=0.01 by chi-square test)

(Table 1).However, the difference for each types of lesions were not statically significant (except CE5, the human sonographer achieved extremely low accuracy). The result were represented as confusion matrix(Figure 3 and Figure 4).

**Table 1.** Comparison of human and AI

|  | CL | CE1 | CE2 | CE3 | CE4 | CE5 | AE1 | AE2 | AE3 | total | acc.* | p-values# |
| --- | --- | --- | --- | --- | --- | --- | --- | --- | --- | --- | --- | --- |
| CL-Human | 2 | 2 | 0 | 0 | 0 | 0 | 0 | 0 | 0 | 4 | 0.50 | 0.27 |
| CL-AI | 38 | 4 | 4 | 0 | 4 | 0 | 0 | 0 | 0 | 50 | 0.76 |  |
| CE1-Human | 1 | 11 | 0 | 0 | 0 | 0 | 0 | 0 | 0 | 12 | 0.92 | 0.68 |
| CE1-AI | 4 | 40 | 4 | 0 | 2 | 0 | 0 | 0 | 0 | 50 | 0.80 |  |
| CE2-Human | 0 | 0 | 2 | 1 | 0 | 0 | 0 | 0 | 0 | 3 | 0.67 | 1.00 |
| CE2-AI | 0 | 0 | 38 | 9 | 3 | 0 | 0 | 0 | 0 | 50 | 0.76 |  |
| CE3-Human | 0 | 0 | 2 | 8 | 1 | 0 | 0 | 0 | 0 | 11 | 0.73 | 1.00 |
| CE3-AI | 0 | 0 | 6 | 38 | 6 | 0 | 0 | 0 | 0 | 50 | 0.76 |  |
| CE4-Human | 0 | 0 | 0 | 1 | 14 | 7 | 0 | 0 | 0 | 22 | 0.64 | 0.15 |
| CE4-AI | 0 | 0 | 8 | 2 | 40 | 0 | 0 | 0 | 0 | 50 | 0.80 |  |
| CE5-Human | 1 | 0 | 0 | 0 | 2 | 0 | 1 | 0 | 0 | 4 | 0.00 | 0.00 |
| CE5-AI | 0 | 1 | 0 | 0 | 1 | 40 | 8 | 0 | 0 | 50 | 0.80 |  |
| AE1-Human | 0 | 0 | 0 | 0 | 2 | 0 | 19 | 0 | 0 | 21 | 0.90 | 0.21 |
| AE1-AI | 0 | 0 | 0 | 0 | 0 | 0 | 49 | 0 | 1 | 50 | 0.98 |  |
| AE2-Human | 0 | 0 | 0 | 0 | 0 | 0 | 1 | 3 | 0 | 4 | 0.75 | 1.00 |
| AE2-AI | 0 | 2 | 0 | 0 | 0 | 0 | 2 | 10 | 0 | 14 | 0.71 |  |
| AE3-Human | 0 | 0 | 0 | 0 | 0 | 1 | 1 | 0 | 2 | 4 | 0.50 | 0.55 |
| AE3-AI | 0 | 0 | 0 | 0 | 0 | 2 | 1 | 0 | 9 | 12 | 0.75 |  |

\* acc. accuracy=right classification/total samples

**Figure 3.**
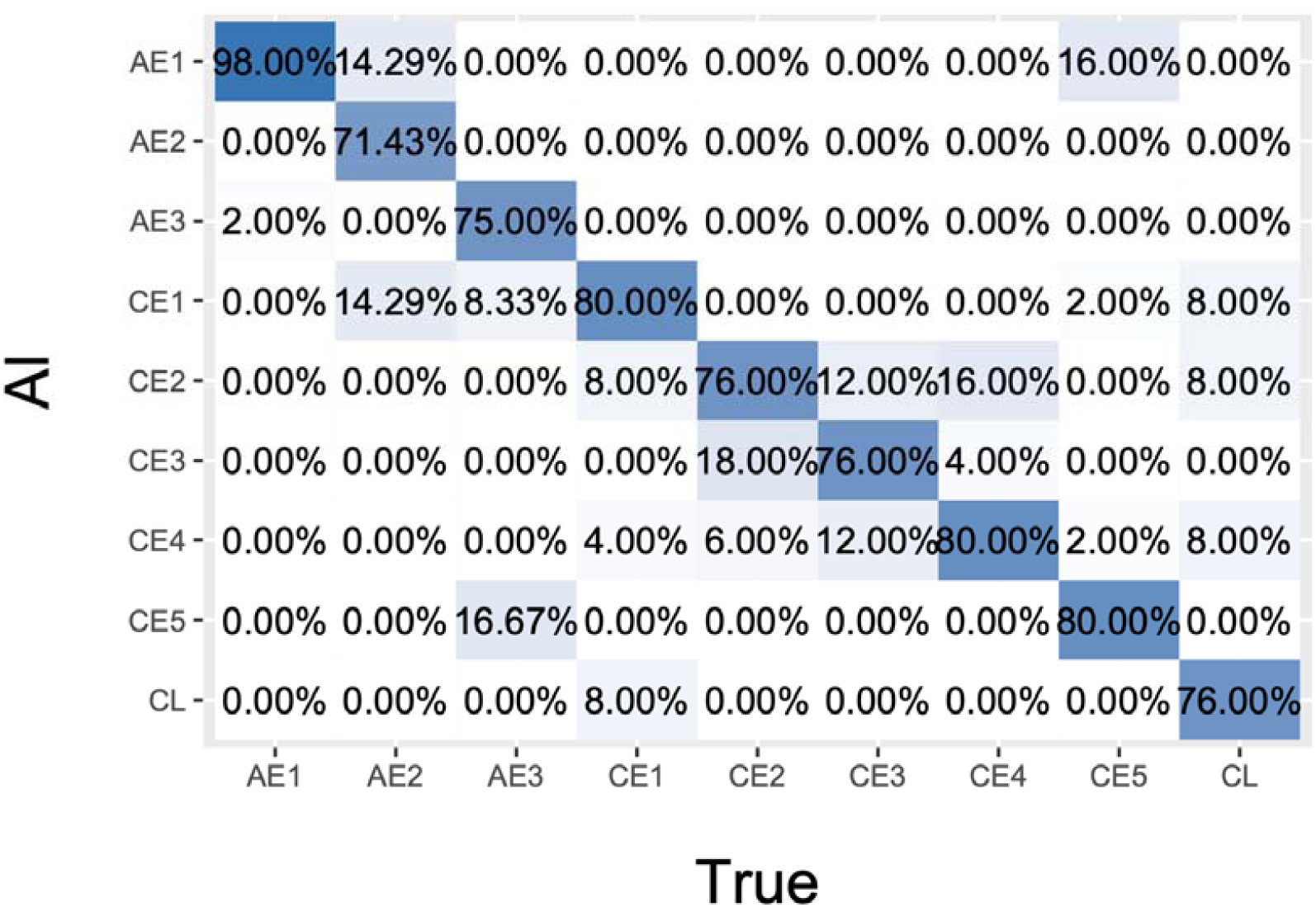
The confusion matrix of AI prediction.

**Figure 4.**
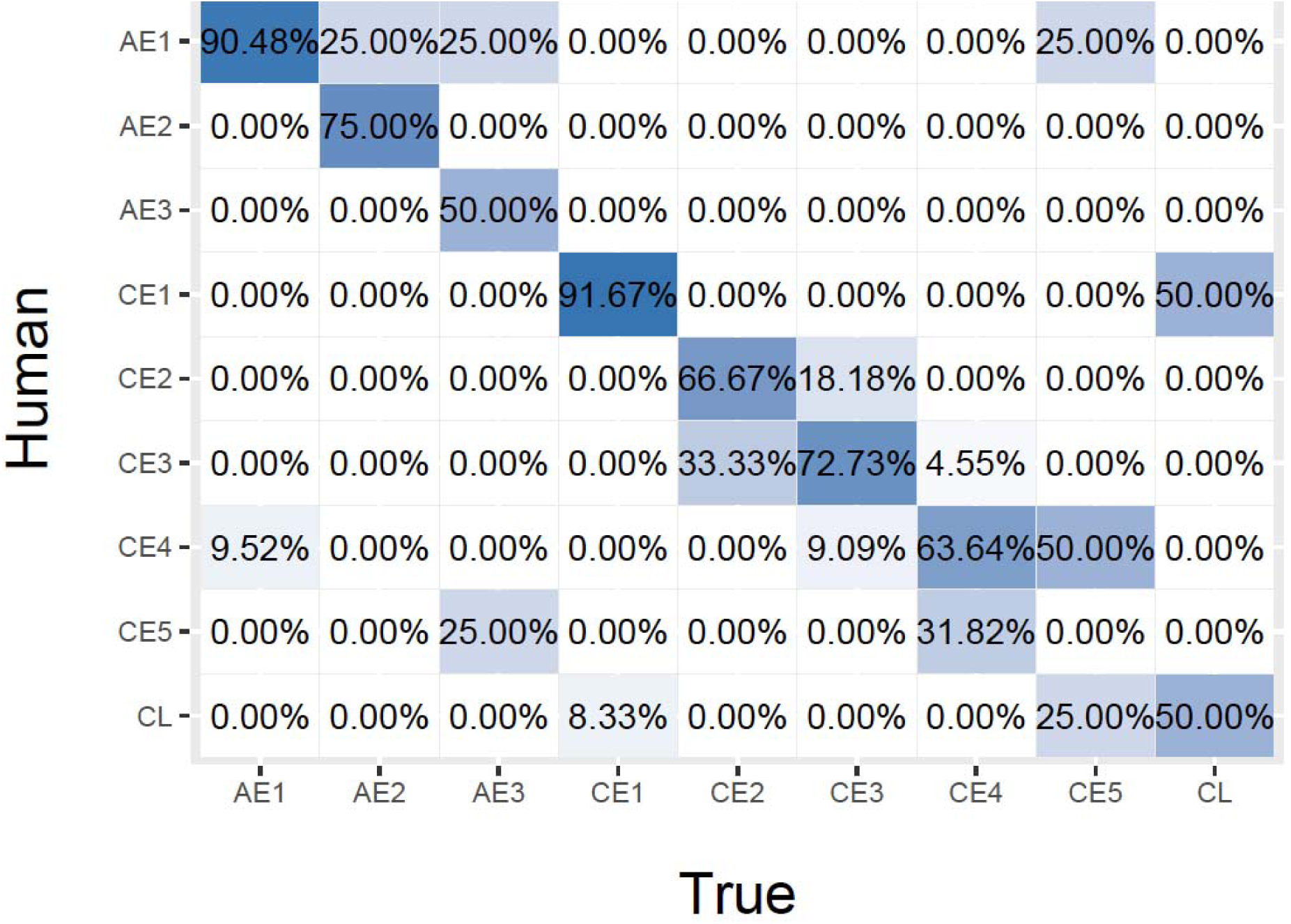
The confusion matrix of human sonographer.

## Discussion

### Model outperforms trained sonographers

According to our testing, our deep learning model outperformed fast-trained sonographer in echinococcosis classification task. The U-net based model has been proven to be efficient for smaller size of training set. It could be highly useful for local paramedics while managing echinococcosis patients.

### Background of the disease

Echinococcus *granulosus* life cycle involves dogs and other canids as definitive hosts for the intestinal tapeworm as intermediate hosts for metacestode (larval) stage. The metacestode (echinococcal cyst). In humans, the slowly growing hydatid cysts can attain a volume of several liters and contain many thousands of protoscolices^31^. The life cycle of E. *multilocularis* involves small rodent intermediate hosts, such as arvicolids, wild or domestic canid definitive hosts, such as red or arctic foxes, jackals, wolves, or dogs. Humans are aberrant intermediate hosts acquiring the infection through ingestion of eggs shed in the feces of definitive hosts. AE is of increasing concern globally due to the geographical spread of the parasite, its increasing prevalence in animals from endemic areas, the absence of a vaccine, and the lack of active control measures to prevent the infection^32^.

### Web-based application improved accessibility of AI tools

Deep learning applications are provided as source code deposited on Github^33^ or similar platform for sharing. It is not feasible for many medical related applications, since the privacy concern of the data. The run-time environments of the source code were varied in wide range due to different platforms, different frameworks, and versions used. Here we used open-access web-based tool available for PC and mobile will significantly increase the accessibility for many remote areas.

### Clinical implications

We demonstrated the AI model achieved better performance than sonographer, it could help doctors in remoted area for echinococcosis lesion typing. The lesion typing for echinococcosis was important for disease managements.

According to the consensus on cystic echinococcosis diagnosis and treatment, five types of lesions were recommended for classification. A natural grouping of the cysts into three relevant groups: active (CE1 and 2), transitional (CE3) and inactive (CE4 and 5). The lesion classification is critical to patients managements, according to the consensus, CE4 and 5 indicated an inactive infection, watch and wait was recommended, if patients with CE1-3, the consensus suggested an active larva infection, radical medical surgery or/and ABZ should be suggested^11,2^.

Comparing CE, AE is more problematic, surgical resection, chemotherapy, early diagnosis, and multidisciplinary discussion contributed to the successful treatment for AE cases^11^. AE lesions behave “cancer-like”^34^, radical surgery is the first choice in all cases suitable for totally resection^11^. Benzimidazoles should be used for all cases^11^, and liver transplantation as an alternative to palliative surgery, however, has not been shown to be superior to long-term conservative therapy^35^.

## Data Availability

The data and codes in this study are available from the corresponding author on request.

